# Spread of SARS-CoV-2 Coronavirus likely constrained by climate

**DOI:** 10.1101/2020.03.12.20034728

**Authors:** Miguel B. Araújo, Babak Naimi

**Affiliations:** Department of Biogeography and Global Change, National Museum of Natural Sciences, CSIC, Calle Jose Gutierrez Abascal, 2, 28006, Madrid, Spain; Rui Nabeiro Biodiversity Chair, MED Institute, University of Évora, Largo dos Colegiais, 7000, Évora, Portugal; Department of Geosciences and Geography, University of Helsinki, 00014, PO Box 64, Helsinki, Finland

## Abstract

As new cases of COVID-19 are being confirmed pressure is mounting to increase understanding of the factors underlying the spread the disease. Using data on local transmissions until the 23rd of March 2020, we develop an ensemble of 200 ecological niche models to project monthly variation in climate suitability for spread of SARS-CoV-2 throughout a typical climatological year. Although cases of COVID-19 are reported all over the world, most outbreaks display a pattern of clustering in relatively cool and dry areas. The predecessor SARS-CoV-1 was linked to similar climate conditions. Should the spread of SARS CoV-2 continue to follow current trends, asynchronous seasonal global outbreaks could be expected. According to the models, temperate warm and cold climates are more favorable to spread of the virus, whereas arid and tropical climates are less favorable. However, model uncertainties are still high across much of sub-Saharan Africa, Latin America and South East Asia. While models of epidemic spread utilize human demography and mobility as predictors, climate can also help constrain the virus. This is because the environment can mediate human-to-human transmission of SARS-CoV-2, and unsuitable climates can cause the virus to destabilize quickly, hence reducing its capacity to become epidemic.

## Introduction

Biogeography studies the patterns and processes underlying the distribution of Life on earth. One generalization emerging from more than two hundreds of years of natural history observations is that all organisms have a degree of environmental specialization. That is, while biomes in the planet have a range of different types of organisms(*1*), individual types of organisms cannot occur in every biome even when, distant apart as they might be, they converge into playing the same ecological roles within ecosystems(*2*). Biogeographers and ecologists alike resort to the concept of ecological niche(*3-5*) to examine the relationship between the distributions of organisms and other biotic or abiotic factors controlling them. An organism is said to be within its ecological niche if death rates of the organism are lower that birth rates(*6, 7*). That is, an organism cannot persist beyond its ecological niche, in a sink, unless there is a regular influx of individuals from source populations. Even if the organisms are regularly reaching a sink area, as one might expect with an easily dispersed pathogen, the spread and establishment of the organism will be limited by ecological constraints. Although biogeographic concepts, such as species ecological niches, are commonly used and applied to multicellular organisms (eukaryotes), there is an increased number of studies utilizing the ecological concepts and associated analytical tools to investigate relationships between the distributions of unicellular organisms (prokaryotic), or viruses, and a range of biotic and abiotic environmental factors(*8-10*).

Building on the concept of ecological niche, we develop projections of monthly changes in the climate suitability for SARS-CoV-2 outbreaks. Projections are obtained from an ensemble of 10 familiar machine learning ecological niche models(*11*), each with 20 replications generated by repeating four times a 5-fold cross validation that accounts for and enables the quantification of variability across initial conditions(*12*). Models were trained using the distribution of all recorded local transmissions of SARS-CoV-2 Coronavirus (excluding imported cases) with data compiled from John Hopkins University Mapping 2019-nCoV portal(*13*). Predictors were monthly mean temperature, interaction term between monthly minimum temperature and maximum temperature, monthly precipitation sum, downward surface shortwave radiation, and actual evapotranspiration. Climate data refer to a period including January, February and March 2009–2018, with data downloaded from the updated high-resolution global climatology database - TerraClimate(*14*). Models were then projected monthly for the rest of the year.

## Results

Local transmissions of SARS-CoV-2 Coronavirus were plotted against monthly values of climate predictors revealing aggregation within a relatively narrow range of climatic values (Figure 1). Comparing daily spread of the virus across geographical space versus climatic space reveals stronger aggregation in climate space than in geographical space. That is, while the virus is progressively colonizing most parts of the world, thus being geographically widespread, local infections were still prevalent within a relatively narrow set of environmental conditions (Figure 2). The uneven colonization of geographic versus climatic space, invites the interpretation that climate is acting as a stronger constraint for the spread of the virus than geographical distances are. Most local transmissions occur in regions exposed to cool and dry conditions—measured both through evapotranspiration and precipitation—, and near the lower end of the radiation gradient (but *rho* with temperature = 0.78).

**Figure 1.**
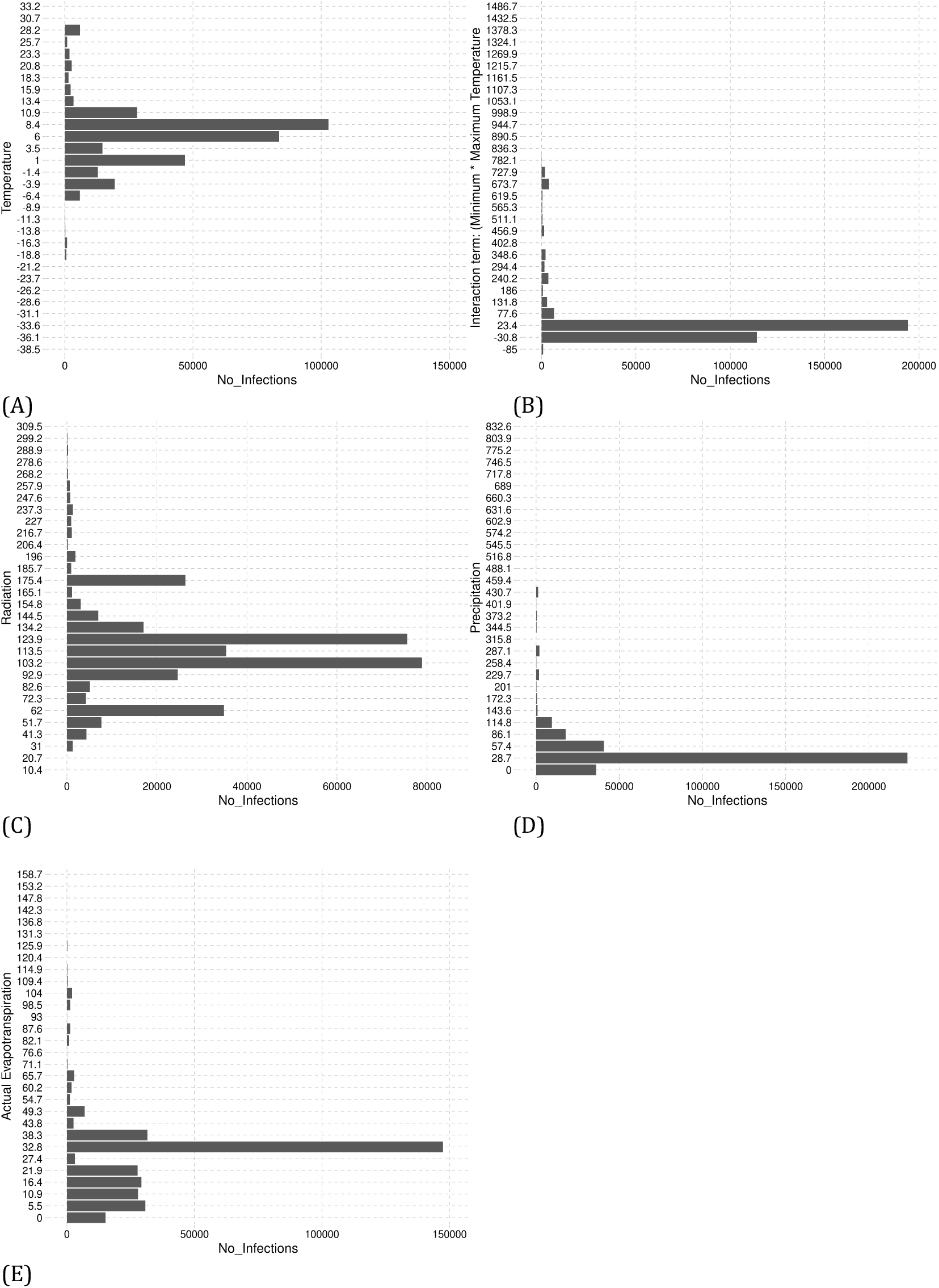
Frequency distribution of COVID-19 positive cases plotted against the world gradient of mean temperature (A), interaction of minimum temperature * maximum temperature (B), downward surface shortwave radiation (C), precipitation (D), and actual evapotranspiration (E) in a typical climatological series between January and March.

**Figure 2.**
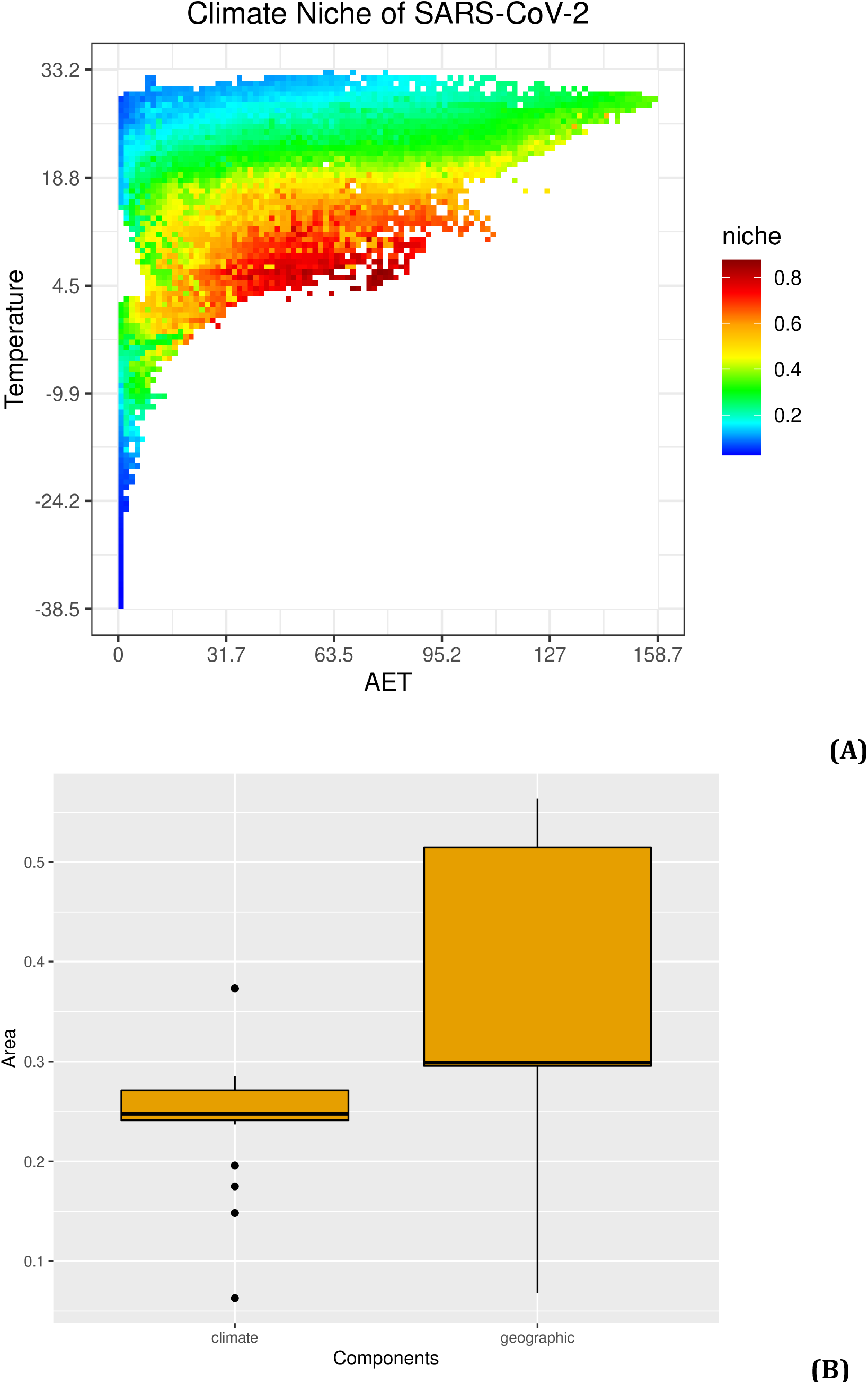
COVID-19 cases in geographical and climate niche space from the 22^nd^ of January to the 23^rd^ of March 2020. (A) Weighted average consensus modeled climate suitability across mean temperature and actual evapotranspiration. (B) Variation of daily convex-hull-polygon area of COVID-19 cases in geographical and climatic space (see methods): the greater area of polygons, the greater the spread. Black lines represent mean values, boxes the 2^nd^ and 3^rd^ interquartile range, lines the 1^st^ and 4^th^ interquartile range, and dots are outliers. A dynamic visualization of the daily spread of SARS-CoV-2 Coronavirus is available here: http://www.maraujolab.com/wp-content/uploads/2020/03/corona_niche_aet-temperature_23march.gif

The mean and interquartile range of average environmental temperatures associated with positive cases is 5,81ºC (mean) and −3,44ºC to 12,55ºC (95% range) and for radiation values are 112,78 W/m^2^ (mean) and 61,07 W/m^2^ and 170,96 W/m^2^ (95% range). For precipitation, the values are 54,73 mm (mean) and 13,6 mm to 115,33 mm (95% range) while for evapotranspiration values are 28,80 mm (mean) and 5,97 mm and 48,44 mm (95% range). These values are estimated taking into account total numbers of positive cases (i.e., abundance), which are obviously strongly determined by contingent factors linked with the origin of the SARS-CoV-2 Coronavirus outbreak (the city of Wuhan in China) and subsequent pattern of spread. While the pattern of spread is likely to be partly constrained by climate, the actual numbers of positive cases are affected by non-climatic factors too(*15*), some of which might be stochastic. Less sensitive measurements can be obtained by using the presence and absence of positive cases. With such an approach, the mean and estimated interquartile ranges for temperature are 9,14ºC (mean) and −11,43ºC to 27,15ºC (95% range), and for radiation they are 154,57 W/m^2^ (mean) and 54,31 W/m^2^ and 255,43 W/m^2^ (95 range). For precipitation they are 71,65 (mean) and 3,19 mm to 236,50 mm (95% range), while being 39,84 mm (mean) and 0,00 mm and 106,75 mm (95% range) for evapotranspiration.

Ecological niche models were used to estimate the combination of climate values best explaining infections with SARS-Cov-2. While the exact response curve to each one of the predictor variables changes slightly with the modeling technique and cross-validation repetition, they are generally consistent amongst themselves (supplementary Figure S1) and with the raw distribution of positive cases across gradients (Figure 1). Mean temperature and evapotranspiration (a surrogate of humidity) are the predictors best explaining the distribution of outbreaks of the virus, with areas either too cold or too hot, or too wet, having lower exposure to outbreaks. These two climatic variables are followed in importance by the interaction between minimum temperature and maximum temperature and radiation (supplementary Figure S2).

Projections by models reveal the existence of likely seasonal changes in climate suitability for SARS-CoV-2 (Figure 3). From April to September, much of higher latitude regions of the southern hemisphere are projected to face increases in climate suitability for outbreaks of SARS-CoV-2. That includes much of southern America, southern Africa and Southern Australia. Models also project that high latitude regions of the northern hemisphere might be badly hit by the virus as temperatures rise during the summer period. Areas exposed to increases in climate suitability for the virus include Canada and Russia, but also the Scandinavian countries. High elevation areas in the Andes and the Himalayas share the same prospects. Concurrently, areas that, as we speak, are of extreme concern in the northern hemisphere (chiefly Italy, Spain, France, Germany, UK, and USA) could witness a reduction in climate suitability for spread of SARS-CoV-2 between June and September. Beyond September and until the end of May, beginning of June, climate conditions are projected to be suitable for renewed outbreaks in much of the warm temperate regions of Asia, Europe and North America.

**Figure 3.**
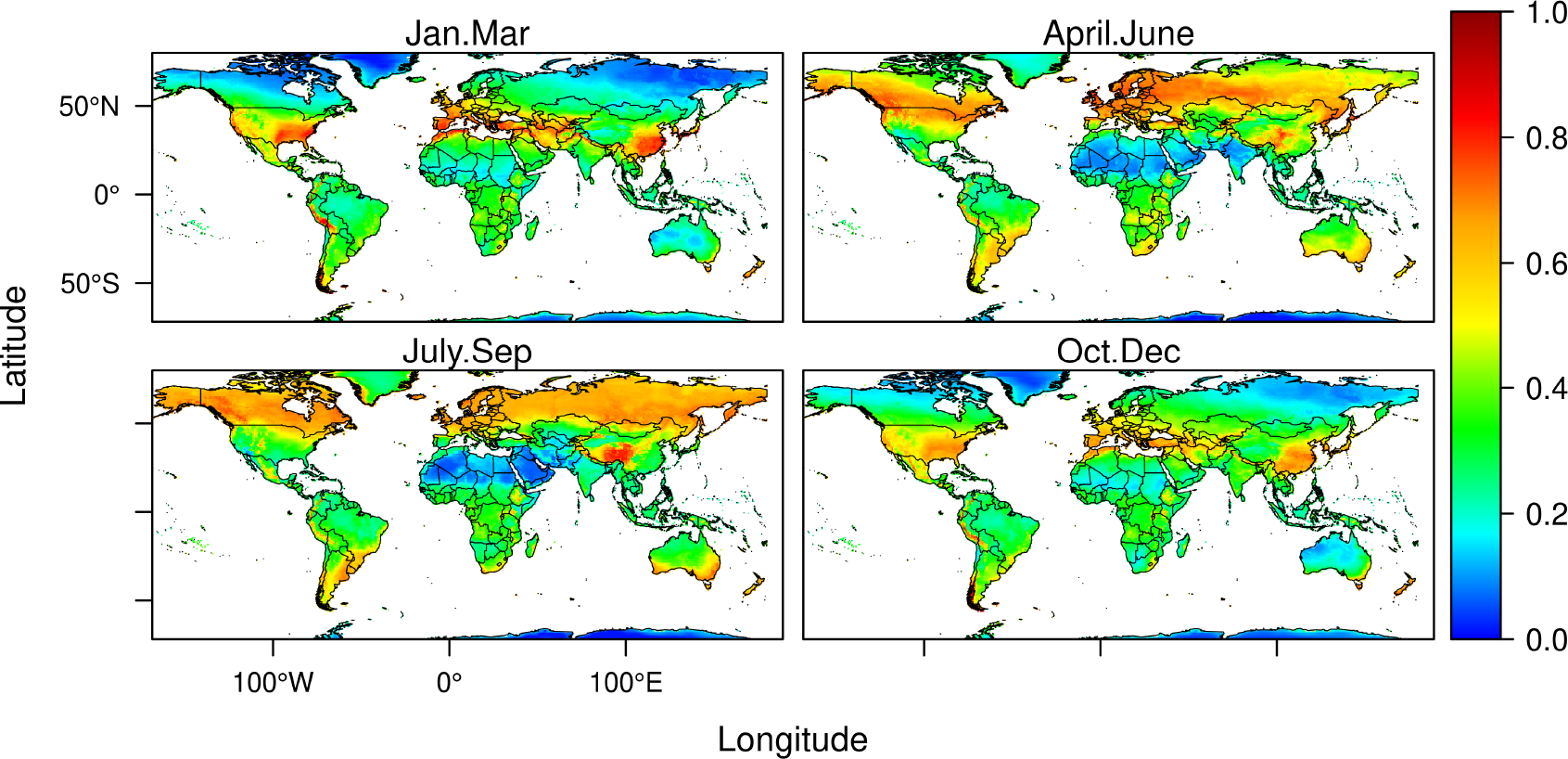
Projected relative climate suitability for SARS-CoV-2 Coronavirus outbreaks across the 5 Köppen–Geiger climate zones of the world(*16*). (A) Distribution of coarse Köppen–Geiger climate zones. (B) Monthly changes in relative climate suitability for the Coronavirus per climate zone. A dynamic visualization of the monthly geographical spread of modeled climate suitability from January to December is available here: http://www.maraujolab.com/wp-content/uploads/2020/03/corona_risk_23march.gif

Projections of seasonal changes in climate suitability can be aggregated across the classic Köppen–Geiger climate zones(*16*) of the world to summarize patterns of climate suitability for the virus (Figure 4). The analysis reveals that climate suitability for SARS-CoV-2 is greater across warm temperate climates from October to April. From April to September cold temperate regions are exposed highly suitable climate conditions for spread of the Coronavirus with a peak of suitability in polar climates between June and August. Arid environments have moderate suitability for the virus, with slight increases from March to April, while the tropics are also moderately suitable with increased climate suitability between June and August.

**Figure 4.**
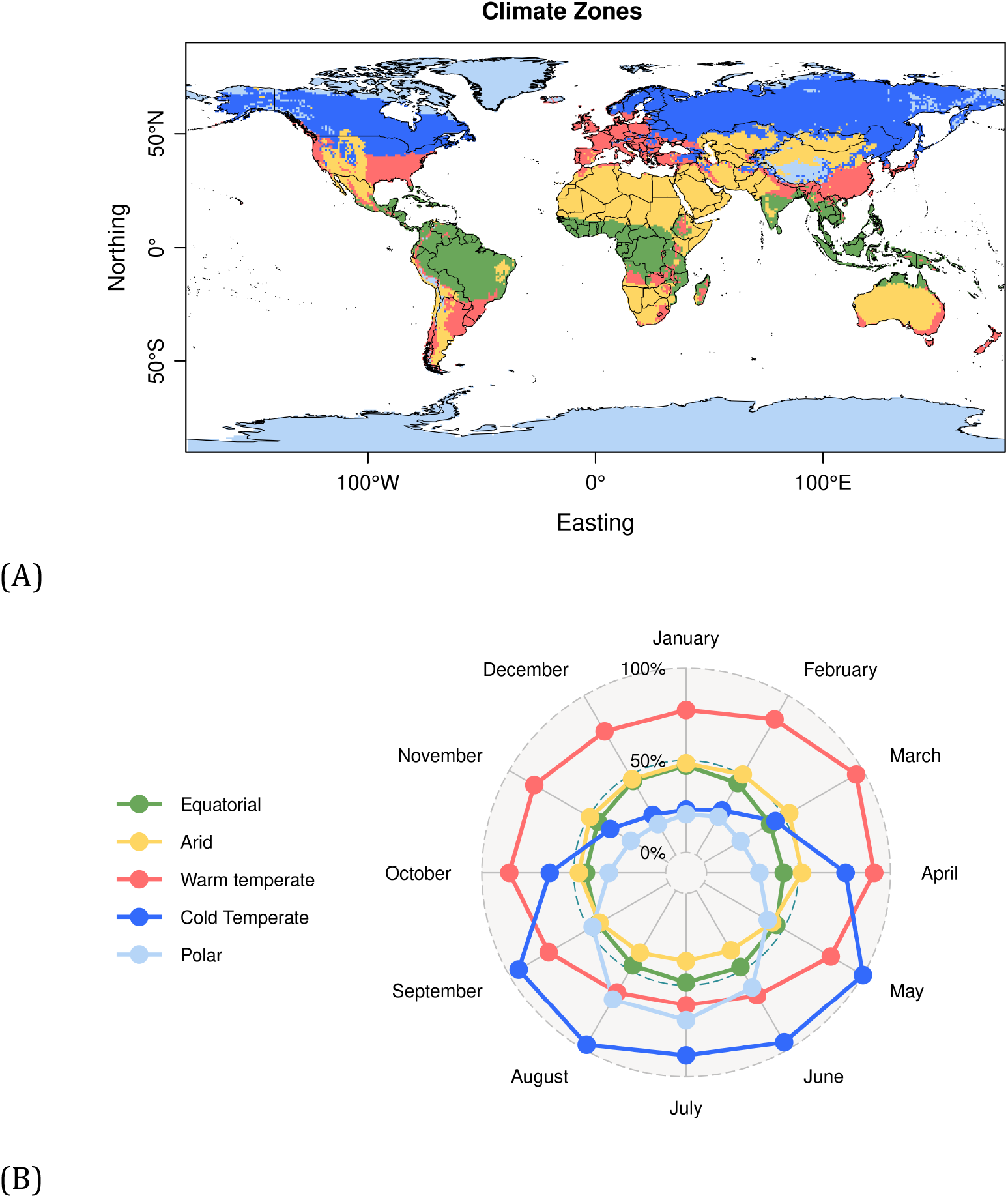
Projected climate suitability for SARS-CoV-2 Coronavirus outbreaks in a typical January-March (A), April-June (B), July-September (C), and October-December (D).

Metrics of model performance on the test data were generally high (mean/SD AUC=0,76/0,03, see supplementary Figure S4). However, models with high performance in the test data can still generate projections that are uncertain when used for forecasting(*17*). While improvements in the data and the models can reduce uncertainties, an issue we are now exploring at light of the recently published standards for applied ENMs(*18*), characterizing the uncertainty of existing models is a first step towards understanding the limitations of projections and highlighting areas of concern. We generated 200 models by varying the initial conditions and model classes. As such, we were able to quantify and map some of the methodological uncertainties associated with projections (Figure 5). Using this approach, we show that variation associated with changing initial conditions is negligible, which is unsurprising given that splits are randomly performed. In contrast, consistently with previous studies addressing climate change forecasts(*19*), uncertainty associated with use of different modeling techniques is high(*20, 21*). Models are considerably variable in areas projected to have low seasonal variability in climate suitability across much of Latin America, sub-Saharan Africa and South East Asia. Specific areas in India, China, Western and Central Europe, Coastal Australia, and Central USA also have high levels of model variability. Such variability is probably a consequence of the sparse nature of the positive cases of COVID-19 in those areas, causing different models to adjust differently to the data. In contrast, inter-model variability across northern higher latitudes is lower and this is likely because most records between January and March are found there. Such areas of low inter-model variability coincide with areas where seasonal variation is also greater.

**Figure 5.**
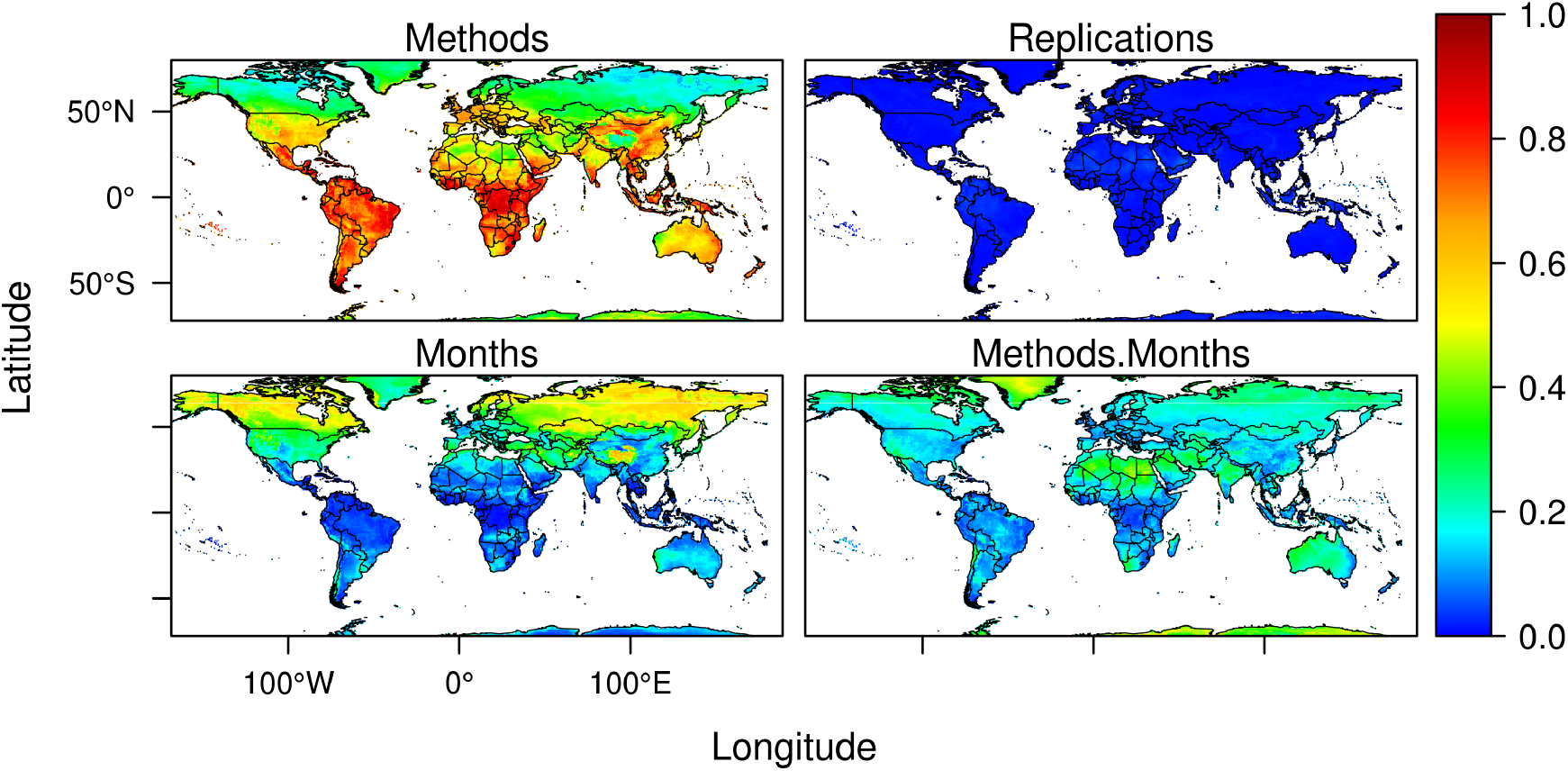
Variation in model projections measured as proportion of the total sum of squares accounted for by ENM methods (A), Cross-Validation replications (B), Monthly seasonal variation (C), and interactions between ENM methods and seasonal monthly variation (D).

## Discussion

Seasonal variation in climate is ubiquitous and it can exert strong pressures on spatial and temporal dynamics of virus-transmitted diseases(*8*). Of course, not all viruses are climate determined. HIV/AIDS, for example, is not affected by external climatic factors. The virus is transmitted by sexual intercourse, blood transfusions, or from mother to child during pregnancy, delivery or breastfeeding, so it never leaves the host’s internal environmental conditions. In contrast, SARS-CoV-2, like other respiratory viruses, namely its predecessor SARS-CoV-1, involves aerial transmissions of respiratory droplets or fomites, exposing the virus to external environmental conditions in which transmissions take place.

SARS-CoV-2 has already set foot in most parts of the world, but virulent outbreaks of COVID-19, with large numbers of local infections, are still clustered in areas with relatively well-defined climate conditions (Figure 2). Starting in Wuhan, China(*22*), the virus quickly became epidemic in several parts of the northern hemisphere, chiefly China, the Middle East, Europe, and the USA. Arguably, being China well connected to the World, the virus would have had equal probability to spread and become epidemic everywhere. But this is not what data show. The pattern of spread, far from being random, was tightly associated with the climate conditions of the temperate and arid zones during the winter. Even the USA that imposed an early ban on flights from China faced an escalation of cases of COVID-19 by the end of March. Our models fitted on the existing pattern of spread between January and March 2020, support the view that incidence of the virus could follow a seasonal climate pattern with outbreaks generally being favored by cool and dry weather, while being slowed down by extreme conditions of both cold and heat as well as moist. Prevalence of respiratory disease outbreaks, such as influenza, during wintering conditions is common(*23, 24*). But the similarity of climate determination of SARS-CoV-2 with its predecessor SARS-CoV-1 and even MERS-CoV is noteworthy, raising reasonable expectations that fundamental traits shared by at least these three Coronavirus might be conserved.

Previous analyses of SARS-CoV-1 outbreaks in relation to meteorology revealed significant correlations between the incidence of positive cases and aspects of the weather. For example, an initial investigation linking SARS outbreaks and temperature in Hong Kong, Guangzhou, Beijing, and Taiyuan(*25*), revealed significant correlations between SARS-CoV-1 incidences and temperature seven days (the known period of incubation of SARS-CoV-1) before the outbreak, with environmental temperatures associated with positive cases of SARS-CoV-1 ranging between 16ºC to 28ºC. They also found that incidence of the Coronavirus was inversely related to humidity. Another study conducted between 11 March and 22 May 2003 in Hong Kong(*26*) showed that SARS-CoV-1 incidences sharply decreased as temperature increased from 15ºC to 29ºC, after which it practically disappeared. In this study, incidences under the cooler end of the gradient were 18-fold higher than under the opposite warmer end of the gradient.

The mechanism underlying these patterns of climate determination is likely linked with the ability of the virus to survive external environmental conditions prior to reaching a host. For example, a recent study examined survival of dried SARS-CoV-1 Coronavirus on smooth surfaces and found that it would be viable for over 5 days at temperatures ranging between 11-25ºC and relative humidity of 40-50%, drastically loosing viability as temperatures and humidity increased(*27*). Likewise, an experiment examining the stability of MERS-CoV on plastic and steel surfaces, under three environmental treatments (20ºC – 40% relative humidity, 30ºC-30% RH, 30ºC – 80% RH), revealed that the virus was more stable in 20ºC and 40% RH treatment, decaying gradually in the second and third treatments(*28*).

Heat intolerance of the Corona viruses is probably related to them being covered by a lipid bilayer(*29, 30*), which could breakdown easily as temperatures increase. Humidity in the air is also expected to affect the transmissibility of respiratory viruses. Once the pathogens have been expelled from the respiratory tract by sneezing, they literally float in the air and they do so for a longer period when the humidity is reduced.

Detailed observational and controlled experiments examining the relationship of SARS-CoV-2 outbreak relationships with weather events are still limited, but there is experimental evidence that SARS CoV-1 and CoV-2 have similar patterns of stability, being viable in aerosols and on surfaces for similar amounts of time(*31*). A recent observational study conducted across 100 cities in China also found that high temperatures and high relative humidity was significantly associated with reductions in reports of COVID-19 cases(*32*). They found their results to be maintained after controlling for population density and GDP (gross domestic product), concluding that the arrival of the summer and rainy season in the northern hemisphere could help reduce the spread of SARS-CoV-2. Another study examined transmission rates of SARS-CoV-2 in the Hubei Province against measurements of absolute humidity(*33*). They found no support for the hypothesis that high humidity would limit survival and transmission of the virus, but the interaction of temperature and humidity on SARS-CoV-2 was not examined.

As the virus spreads and additional climate regions witness outbreaks of COVID-19, it is possible that the climate signal might be weakened or even lost. This could happen as consequence of varying approaches to the management of the epidemics across political units: for example, whether a country choses mitigation versus suppression should effect height and width of the epidemic curve(*34*), hence the frequency distribution of COVID-19 cases in space and time. Likewise, measured relationships between outbreaks of COVID-19 and climate could be contaminated by data with substantially different quality, as it might be the case when comparing regions spending vast amounts of resources in testing for cases and regions practically not any conducting any tests. Countries like Brazil, for example, with a combination of high human population density and absence of proactive political response to COVID-19(*35*), could face epidemic outbreaks even under moderate climate suitability.

Although climate is one of many factors likely affecting the spread of the virus, there is no doubt that the host behavior(*36*) and density(*37*) are powerful predictors of the capacity of the virus to spread. Indeed, projections of climate suitability for the virus are relevant insofar transmissions are made outdoors. Indoor transmissions, under acclimatized environments, can be predominant in certain cultures. That infected humans can be asymptomatic and transmit the virus to other humans, generates substantial uncertainties regarding the overall risk of epidemic outbreak of COVID-19 under a variety of different ecological and social settings(*38*). In China, for example, indoor transmissions were estimated to account for nearly 80% of the total transmissions but this value is likely to change in different cultural and socio-economic contexts. Unfortunately, data regarding the context in which transmissions took place are not readily available.

Understanding the underlying factors involved in the successful spread of SARS-CoV-2 is critical to manage the timing and scale of the social, economic, and political reactions to it. Our results are qualitatively similar with those of two other independent large-scale investigations of the relationship of COVID-19 with geographical and climate factors(*39, 40*). We expect that the SARS-CoV-2 Coronavirus should continue to spread owing to seasonal changes of climate suitability and increased abundance of the viral pool. However, it is unlikely that outbreaks will happen everywhere with the same intensity. Not just because of seasonality in climate, but also because of the interactions with initial date of outbreak, which varies from country to country, and the varying responses to it.

Our results will hopefully contribute to anticipating the timing and magnitude of public interventions needed to mitigate the adverse consequences of the COVID-19 on public health. However, they do not substitute traditional epidemiological modeling and management of the disease that focus on host behavior management as a strategy to minimize overall risk of spread.

## Data Availability

All data is in public repositories.

## Methods

### SAR-CoV-2 Coronavirus data

We downloaded the geo-referenced coordinates of COVID-19 cases from the data repository operated by the Johns Hopkins University Center for Systems Science and Engineering with support from ESRI Living Atlas Team and the Johns Hopkins University Applied Physics Laboratory (https://github.com/CSSEGISandData/COVID-19/blob/master/README.md). The data were downloaded on 23/03/2020. Classification of positive cases into local transmissions and imported cases were obtained from World Health Organization Covid-19 Situation Report(*41*).

### Climate data

We downloaded updated temperature (mean, maximum, minimum), precipitation (accumulated), actual evapotranspiration, and shortwave radiation from “Terra Climate” (a high-resolution global dataset of monthly climate and climatic water balance; http://www.climatologylab.org/terraclimate.html)(*14*). This is a high-resolution (1/24°, ∼4-km) climate and water balance data set for global terrestrial surfaces. The data we downloaded covers the period starting in January 2009 until December 2018, thus covering the recent period of warming that would not be captured by longer climatological time series. The time series data were then averaged, so to provide values for a typical climatological month in the recent past.

### SARS-CoV2-spread across geographic and climate space

This analysis refers to Figure 2B. We used daily data on reported COVID-19 positive cases to calculate a daily convex hull polygon around the coordinates of the positive cases recorded in both geographic and climate space (based on mean temperature and evapotranspiration). To make coordinates comparable in geographic and climate space, coordinate values were re-scaled to a range of 0 to 1. Then, we calculated the area of the polygon in the two spaces each day between the 22^nd^ of January until the 23^rd^ of March: the greater the area of the polygon, the greater the spread of the virus across available geographic and climate space.

### Ecological niche models

The spatial distribution of SAR-CoV-2 Coronavirus records were linked to the corresponding monthly climate data. We used sdm-R platform(*11*) for ensemble ecological niche modeling(*12*) (or species distributions modeling(*42*)), to characterize climate conditions associated with outbreaks of SARS-CoV-2 between January and March 2020. We used 10 commonly used machine learning methods including Generalized Linear Models (GLM)(*43*), Generalized Additive Models (GAM)(*44*), Classification and Regression Trees (CART)(*45*), Boosted Regression Trees (BRT)(*46*), Random Forests (RF)(*47*), Multiple Discriminant Analysis (MDA)(*48*), Multi-Layer Perceptron Neural Networks (MLP)(*49*), Maximum Entropy (Maxent)(*50*), Likelihood-Based Estimator Adopted for Presence-Only Data (Maxlike)(*51*), and Multivariate Adaptive Regression Splines (MARS)(*52*). Models were parameterized using default options of sdm-R as detailed in the Supplementary Material.

We used a 5-fold cross-validation(*53*), repeated 4 times, resulting in 20 replications per method. We then fitted ecological niche models for each replication using the four random cross-validation splits as training set and evaluated them against the fifth withheld data split.

We used the area under curve (AUC) of receiver operating characteristic (ROC) plot and the true skill statistic (TSS) to measure the predictive performance of models(*54*). A ROC curve plots sensitivity values (true positive fraction) on the y-axis against ‘1 – specificity’ values (false positive fraction) for all thresholds on the x-axis. AUC is a threshold-independent metric that varies from 0 to 1 and provides a single measure of model performance. AUC values under 0.5 indicate discrimination worse than expected by chance; a score of 0.5 implies random predictive discrimination; and a score of 1 indicates perfect discrimination. TSS is calculated as “sensitivity + specificity −1” and ranges from −1 to +1, where +1 indicates perfect agreement, a value of 0 implies agreement expected by chance, and a value of less than 0 indicates agreement lower than expected by chance. We then used the ensemble of 200 models to calculate and project a consensus distribution of climate suitability for the spread of SARS-CoV-2, for each month, across the globe. Consensus was achieved through AUC-weighted mean across all models(*21*). The assessments of model performance on cross-validated samples are reported in Supplementary Figure S3.

### Predictor variables importance

Variable importance(*55*) and response curves(*56*) were estimated to infer the explanatory power and shape of the relationship for each one of the predictor variables used and the distribution of positive cases of SARS-CoV-2. Response curves are generated with the evaluation strip procedure(*56*), implemented in sdm-R(*11*). For each climate variable, the method generates the predicted probability of occurrence over all values in the gradient of the climate variable while the other climate variables are kept at their mean values. Then, visualizing the predicted values against the climate gradient represents the response of the species to the climate variable.

Additional model-independent techniques were also implemented to evaluate the relative variable importance. We assessed the relative contribution of variables to explain the distributions of positive cases of SARS-CoV-2 in models using the variable importance (VI) analysis in sdm-R(*11*). This method is a randomization procedure that measures the correlation between the predicted values of a model given the original predictors, and predictions of the same model but given the perturbed dataset in which the variable under investigation is randomly permutated. If the contribution of a variable to the model is high, then it is expected that the permutation would affect the prediction, and consequently, the correlation is low. Using this approach, ‘1 – correlation’ is considered as a measure of variable importance(*57*).

We assessed the relative contribution of variables to explain the distributions of positive cases of SARS-CoV-2 in models using the variable importance (VI) analysis in. This is a permutation method that measures the relative importance of each predictor variable. This method quantifies the correlation between the predicted values and predictions where the variable under investigation is randomly permutated(*11*). If the contribution of a variable to an SDM is high, then the permutation would affect the prediction values, and consequently, the correlation coefficient would be low. Therefore, ‘1 – correlation’ can be considered as a measure of variable importance.

### Decomposition of sources of variation

We used a 3-way ANOVA to decompose variation in model projections from the 10 different modeling techniques used and 20 replications of the initial conditions (data on SARS-CoV-2 cases). We compared variation arising from data and models with variation associated with projected seasonal changes in climate suitability. The latter is the desired projection, rather than a methodological uncertainty. However, comparison of seasonal projected variation in climate suitability with variation arising from different partitions of data and model classes provides a benchmark against which to compare model variability. If data and model variability was greater than projected seasonal variability, then reliance on the models could be questioned. The analysis involved running a three-way Analysis of Variance (ANOVA) without replication(*58, 59*) for each cell, using climate suitability for SARS-CoV-2 as response variable and ecological niche models (ENM), bootstrapped samples (BS), and months (M) as factors. We then obtained the sum of squares to each of these sources and their interaction (ENM × BS, ENM × M, BS × M, ENM × BS × M). We estimated the variance components as the proportions of the sums of squares for the three sources of variation (and their interaction) with regards to the total sum of squares(*20*). Analyses were performed for each cell in the world grid and we mapped each variance component separately (interaction term was not significant, hence not reported). Monthly variation in climate suitability is the expected outcome of the models. Variability associated with ENM and BS is expected to represent the variability associated with changing initial conditions (BS) and model classes (ENM). They are interpreted as uncertainty estimates and should ideally be much proportionally smaller than monthly variation.

## Acknowledgements

The authors thank and congratulate the John Hopkins University for real-time compilation and release of incidence data for SAR-CoV-2 Coronavirus, without which this investigation would not have been possible. Thanks also to the innumerous peers that made comments and suggestions that led to improvements of the manuscript.

## Author contributions

MBA conceived the study and wrote the manuscript. BN conducted the analysis and prepared graphical material.

## Author’s information

### Competing interests

None to declare.

**Figure S1.**
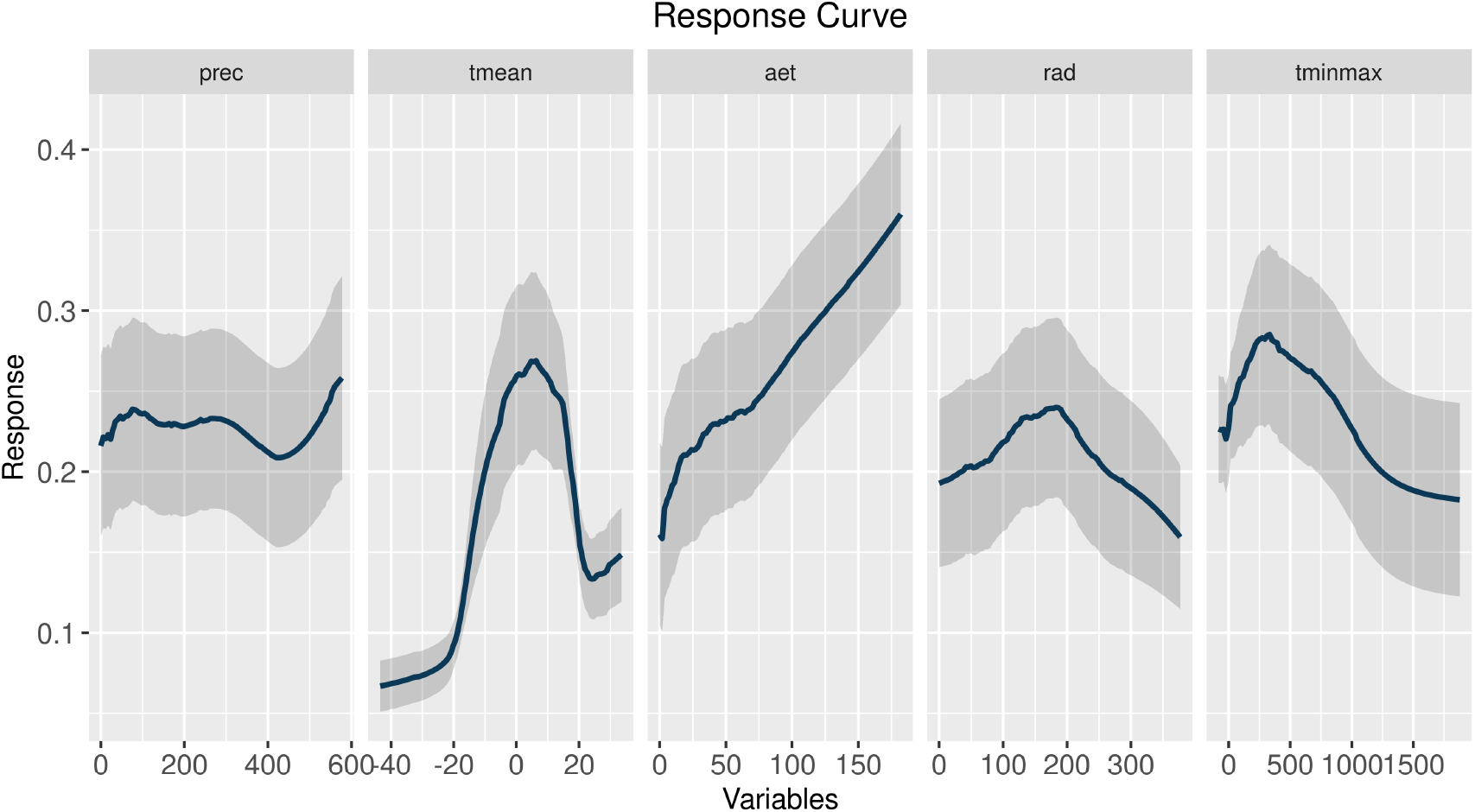
Mean response curves across the 200 ecological niche models of COVID-19 to precipitation, mean temperature, actual evapotranspiration, downward surface shortwave radiation, and interaction between minimum temperature and maximum temperature. Shaded areas represents the 95% confidence interval. See methods.

**Figure S2.**
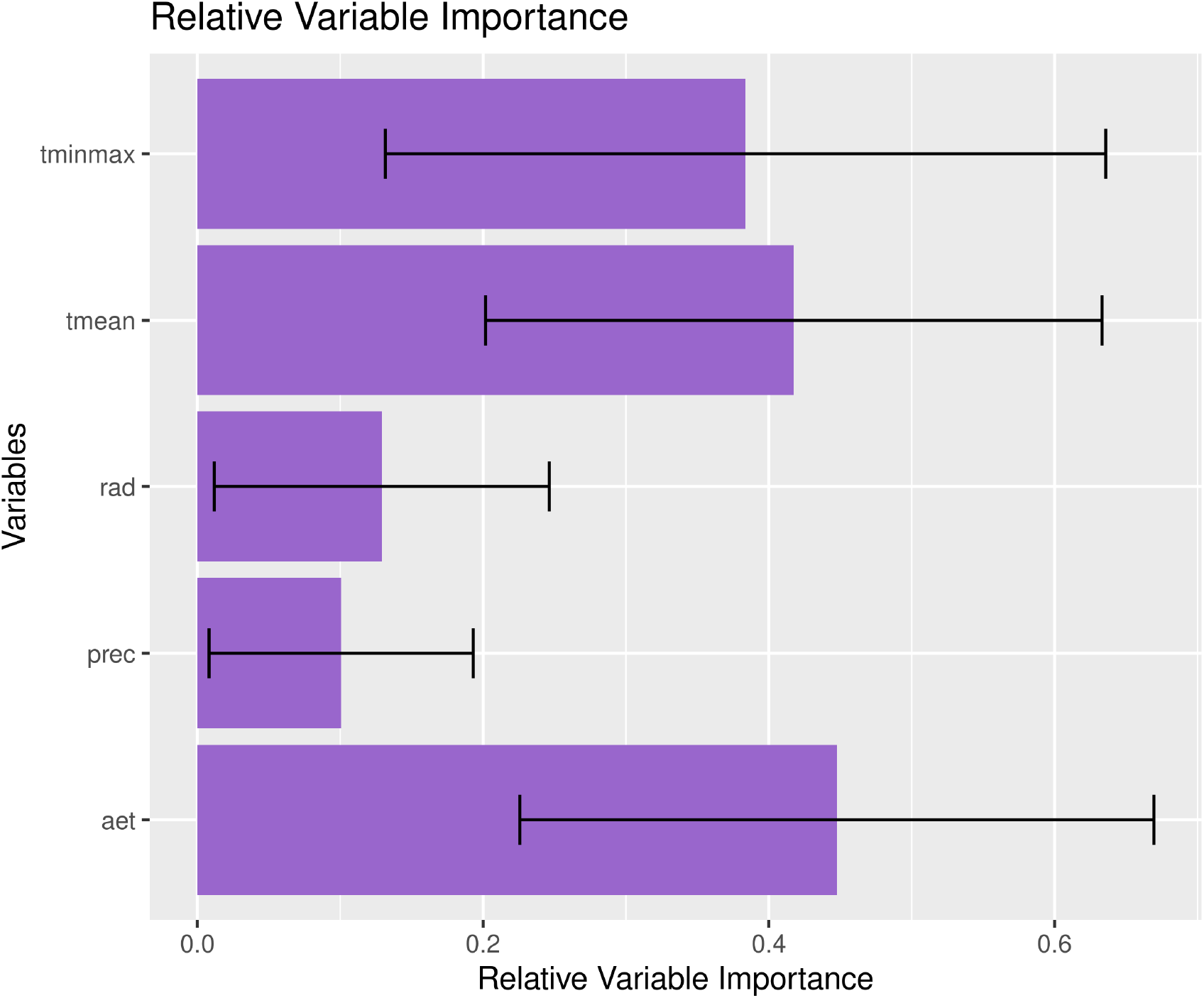
Model-independent estimated relative variable importance of COVID-19 cases. See methods.

**Figure S3.**
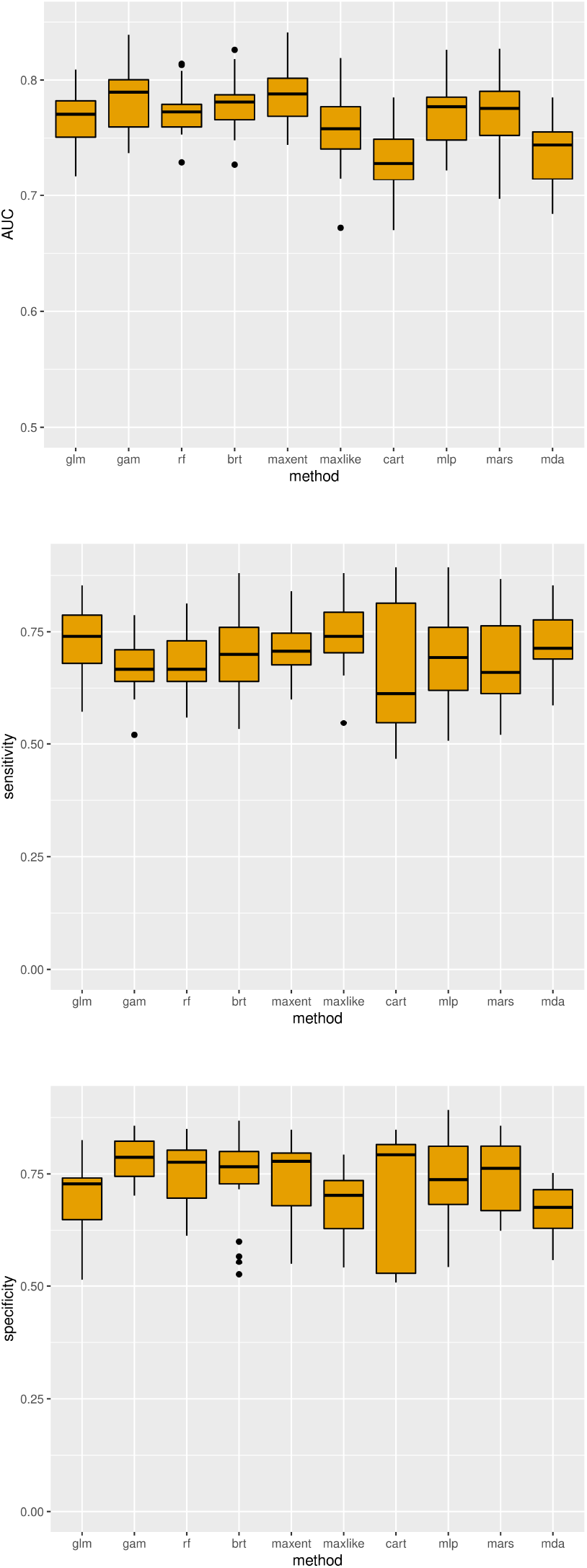
Metrics of performance on test data obtained by 5-fold cross-validation repeated 4 times: AUC; Sensitivity; and Specificity. Black lines represent mean values, boxes the 2^nd^ and 3^rd^ interquartile range, lines the 1^st^ and 4^th^ interquartile range, and dots are outliers.

